# Uncorrected cardiac malformation is the major risk factor for death in patients with dextrocardia

**DOI:** 10.1101/2021.02.05.21251206

**Authors:** Xiaofeng Lu, Wenzhu Dang, Shenglin Li, Xu Wang, Bingren Gao, Debin Liu

**Author notes:** Corresponding author address: Department of Cardiac Surgery, Lanzhou University Second Hospital, No. 82, Cuiyingmen, Chengguan District, Lanzhou, Gansu, 730030, China. **CONFLICT OF INTERESTS** No conflicts of interest or disclosures was declared. Our manuscript has not been previously published or being concurrently under consideration to publish elsewhere. **FUNDING SOURCES** This work was support by Cui Ying Technology Innovation Project of Lanzhou University Second Hospital (CY2017-BJ01) and Lanzhou Talent Innovation and Entrepreneurship Project (2017-RC-63).

## Abstract

**Background/Objectives:** Dextrocardia is rare in the general population. We aimed to analyze the clinical characteristics and risk factors for survival rate of dextrocardia patients during 2013 to 2018 in our hospital, through a specialized Hospital Information System.

**Methods:** A retrospective study of patients with dextrocardia was performed through Hospital Information System. International Classification of Diseases 10 was searched to identify eligible cases from January 2013 to December 2018 in our hospital. Medical records were reviewed to acquire the basic information of dextrocardia patients.

**Results:** Among 9304 patients diagnosed with congenital heart disease in past six years, 48 (0.51%) had dextrocardia. Among 48 dextrocardia patients, 28 (58.33%) had situs inversus viscerum, 20 (41.67%) had cardiac malformation, nine (18.75%) had other malformations and 16 (33.33%) had no malformation. Compared with patients with uncorrected cardiac malformation, patients without cardiac malformation were older (43.5000 vs 0.4583 years, P<0.001). The dextrocardia patients’ age of onset correlated significantly with cardiac malformation. Multivariable logistic model showed the significant risk factor for survival rate was uncorrected cardiac malformation (odds ratio, 22.2391; P <0.001). However, situs inversus (odds ratio, 1.9750; P= 0.4399) or pneumonia (odds ratio, 4.2610; P= 0.0849) itself was not a significant risk factor for survival rate. Kaplan–Meier plot shown that dextrocardia patients with uncorrected cardiac malformation had lower survival rate than those without cardiac malformation (P<0.0001), and there is no significant difference between healthy cohort and patients with pure dextrocardia with or without situs inversus (P=0.25).

**Conclusions:** Dextrocardia remains a rare finding in population, even in the group of patients with known congenital heart disease. Dextrocardia was associated with various malformations besides cardiac malformation. Uncorrected cardiac malformation, which should be paid high attention in the developing countries, is the main risk factor for death in patients with dextrocardia, and pure dextrocardia with or without situs inversus has no effect on survival rate of dextrocardia patients.

## 1 Introduction

Dextrocardia is the most common cause of a congenital cardiac malposition. The incidence which was reported in high-income countries and low-income countries ranges from 0.01% to 0.35% [1-3]. Dextrocardia may be associated with cardiac malformation or extra-cardiac malformation, or without any malformation [4]. Dextrocardia was usually diagnosed in early childhood, and most patients were associated with heart failure and respiratory infections.

The rate of cardiac surgery in dextrocardia patients varied from 23.15% to 53.09% between developing countries and developed countries [1,5]. In the past six years, there were 48 patients with dextrocardia in our hospital. As a remote area in northwest China, only seven (10.29%) patients had cardiac surgery in our retrospective study. And we mainly analyzed the prevalence, clinical features and risk factors for survival rate of patients with dextrocardia.

## 2 Methods

### 2.1 Ethics statement

As a retrospective study, the Ethics Committee of Lanzhou University Second Hospital approved our study. We obtained informed consent from all patients’ family members by telephone follow-up.

### 2.2 Clinical data collection

Based on the latest ICD-10, we identified 205 congenital heart disease diagnostic codes. We acquired 9304 records of patients’ basic information with congenital heart disease through HIS database, including gender, age, ethnicity, visiting time, address, visiting numbers, diagnosis. The first treatment information was included in the analysis when several visiting records were found. A total of 53 cases, 48 patients were diagnosed with dextrocardia. All dextrocardia patients included at least one of the chest X-ray, echocardiography or cardiac computerized tomography angiography. In addition, a healthy cohort was acquired from physical center, with the same sex, age and visiting year as the cohort of dextrocardia without cardiac malformation. Prognosis was obtained through outpatient follow-up and telephone follow-up.

### 2.3 Statistical analysis

Continuous variables are expressed as mean ± standard deviation for normally distributed data and as median (P25, P75) for data with a skewed distribution. Ranked data is described as percentages. The unpaired Student’s t-test, Chi-squared test, Fisher’s exact test or Wilcoxon rank-sum test were employed, as appropriate. Multivariate Spearman correlation was used to find related factors. Multivariate logistic regression analysis was used for find risk factors. Survival analysis was shown by means of Kaplan–Meier plots, and log-rank test was used to compare the survival among uncorrected cardiac malformation group, non-cardiac malformation group and healthy group. A p-value < 0.05 was used as the level of significance. Statistical analysis was performed using R version 3.6.0 (https://www.r-project.org).

## 3 Results

### 3.1 Demographic information

Among the 9304 patients,48(0.516%) patients had dextrocardia, and female to male ratio was 1:1(24/24). The age distribution of 48 patients with dextrocardia is as follows (Figure 1a & 1b). Patients with cardiac malformation were younger than those with no cardiac malformation, and situs inversus made no sense between two groups.

**Figure 1a.**
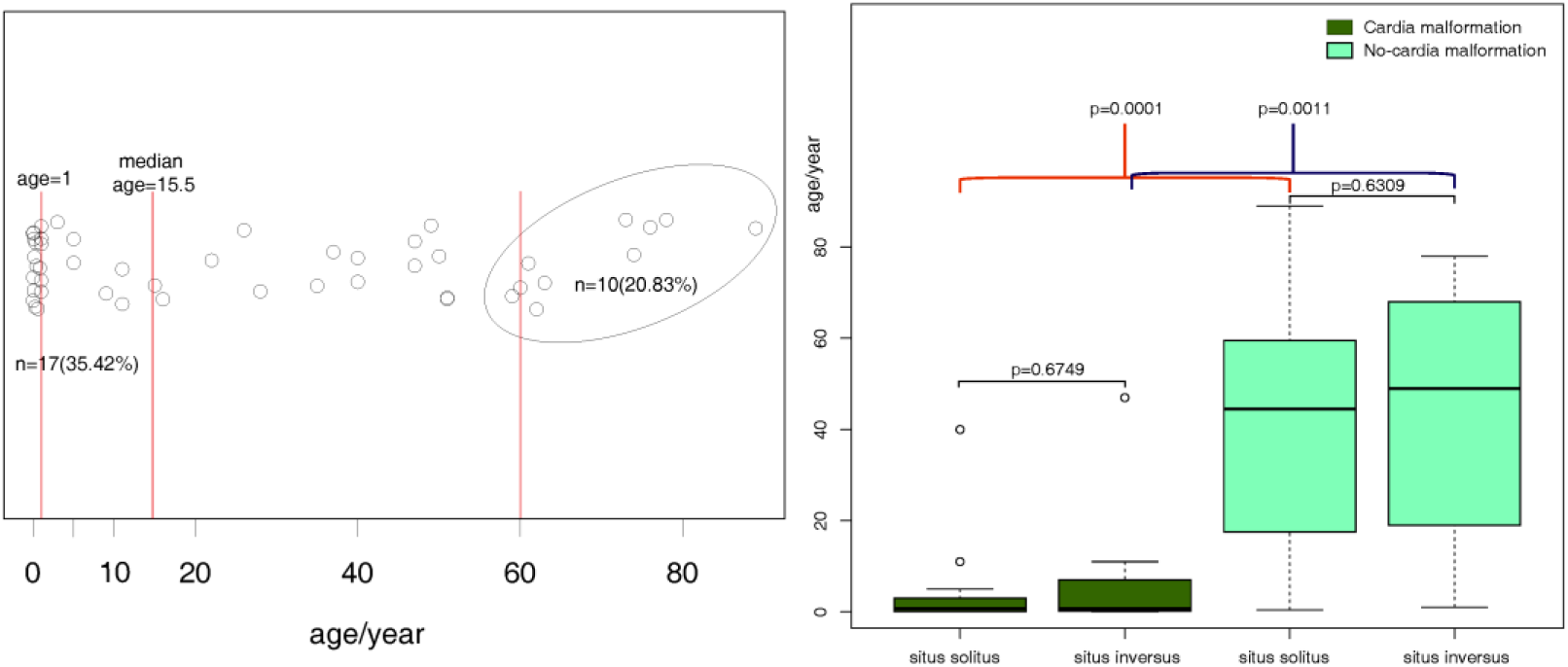
Scatter plot of 48 dextrocardia patients’ age distribution; Figure 1b. Box plots of 48 dextrocardia patients’ age.

### 3.2 Clinical characteristics of patients

Detailed information regarding 48 dextrocardia patients is presented as follows. Among 48 patients with dextrocardia, the most common chief complains were abdominal pain (16.67%) and cyanosis (10.42%). 20(41.67%) patients had respiratory infections, five (10.42%) patients had gallstones and three (6.25%) had arrhythmia.

### 3.3 Risk factor for death in patients with dextrocardia

#### 3.3.1 Baseline characteristics

Baseline characteristics of 48 patients with dextrocardia, which were divided into two groups (dead group & alive group), are presented in Table 1. The age of the patients in alive group was older than that of those in dead group (P = 0.0013). And the follow-up time was longer in the alive group compared to dead group (P<0.001). Uncorrected cardiac malformation was more common in dead group (P<0.001). The proportion of patients with pneumonia was higher in dead group.

**Table 1.**
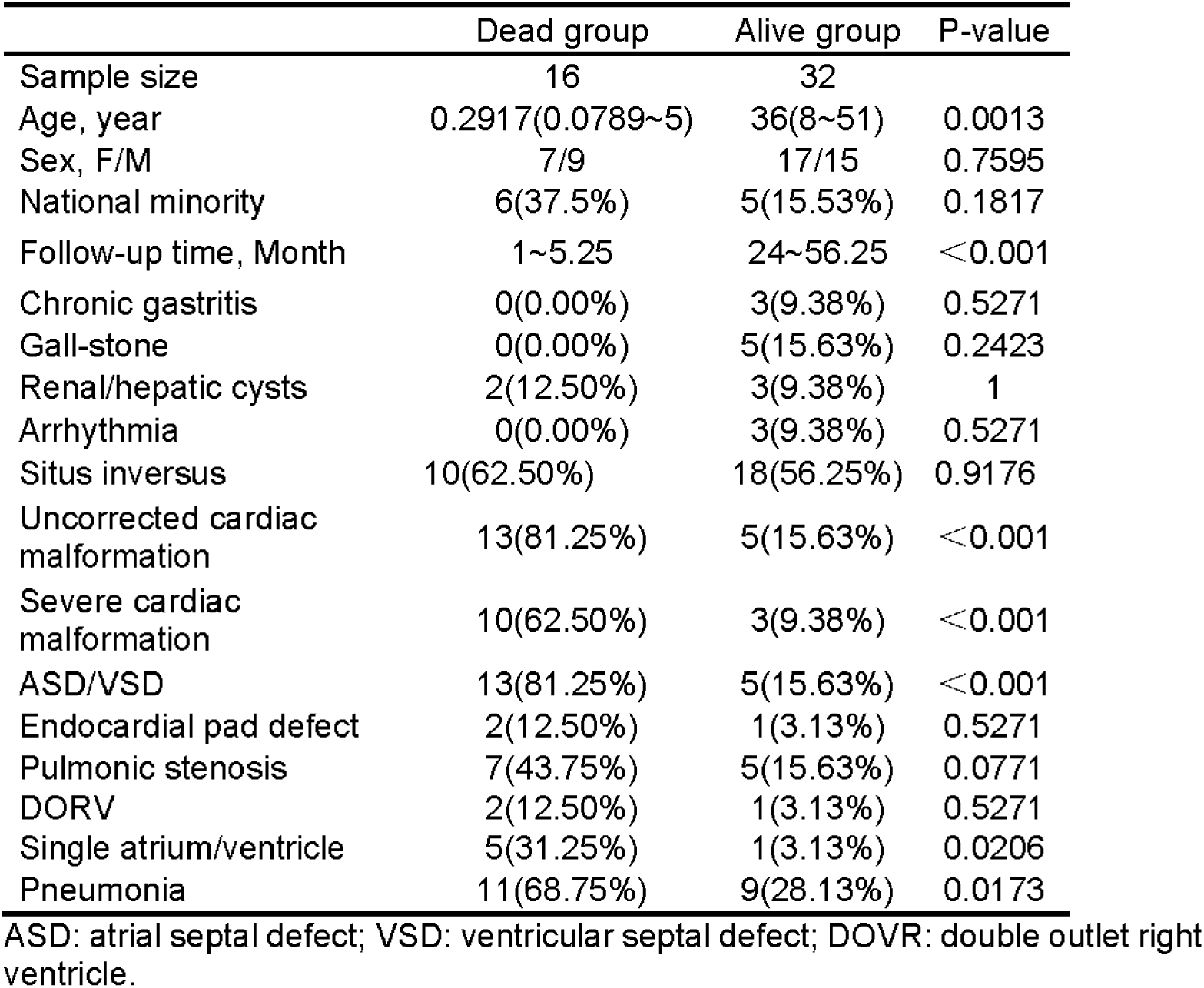
Clinical Characteristics of dextrocardia patients

|  | Dead group | Alive group | P-value |
| --- | --- | --- | --- |
| Sample size | 16 | 32 |  |
| Age, year | 0.2917(0.0789~5) | 36(8~51) | 0.0013 |
| Sex, F/M | 7/9 | 17/15 | 0.7595 |
| National minority | 6(37.5%) | 5(15.53%) | 0.1817 |
| Follow-up time, Month | 1~5.25 | 24~56.25 | <0.001 |
| Chronic gastritis | 0(0.00%) | 3(9.38%) | 0.5271 |
| Gall-stone | 0(0.00%) | 5(15.63%) | 0.2423 |
| Renal/hepatic cysts | 2(12.50%) | 3(9.38%) | 1 |
| Arrhythmia | 0(0.00%) | 3(9.38%) | 0.5271 |
| Situs inversus | 10(62.50%) | 18(56.25%) | 0.9176 |
| Uncorrected cardiac malformation | 13(81.25%) | 5(15.63%) | <0.001 |
| Severe cardiac malformation | 10(62.50%) | 3(9.38%) | <0.001 |
| ASD/VSD | 13(81.25%) | 5(15.63%) | <0.001 |
| Endocardial pad defect | 2(12.50%) | 1(3.13%) | 0.5271 |
| Pulmonic stenosis | 7(43.75%) | 5(15.63%) | 0.0771 |
| DORV | 2(12.50%) | 1(3.13%) | 0.5271 |
| Single atrium/ventricle | 5(31.25%) | 1(3.13%) | 0.0206 |
| Pneumonia | 11(68.75%) | 9(28.13%) | 0.0173 |

ASD: atrial septal defect; VSD: ventricular septal defect; DOVR: double outlet right ventricle.

#### 3.3.2 Correlation analysis

The pairwise correlations among 17 factors are shown in Figure 2. The onset age was also strongly correlated with ASD/VSD, uncorrected cardiac malformation and severe cardiac malformation(no less than three types of cardiac malformation) (r>0.5, P<0.001). The survival rate had positive correlation with follow-up time (r=0.74, P< 0.001) but had negative correlation with cardiac malformation besides ASD/VSD (r=-0.64, P<0.001). ASD/VSD and pulmonic stenosis were common cardiac malformation. As a result, we could draw a conclusion that cardiac malformation was associated with younger onset age, shorter follow-up time and lower survival rate.

**Figure 2.**
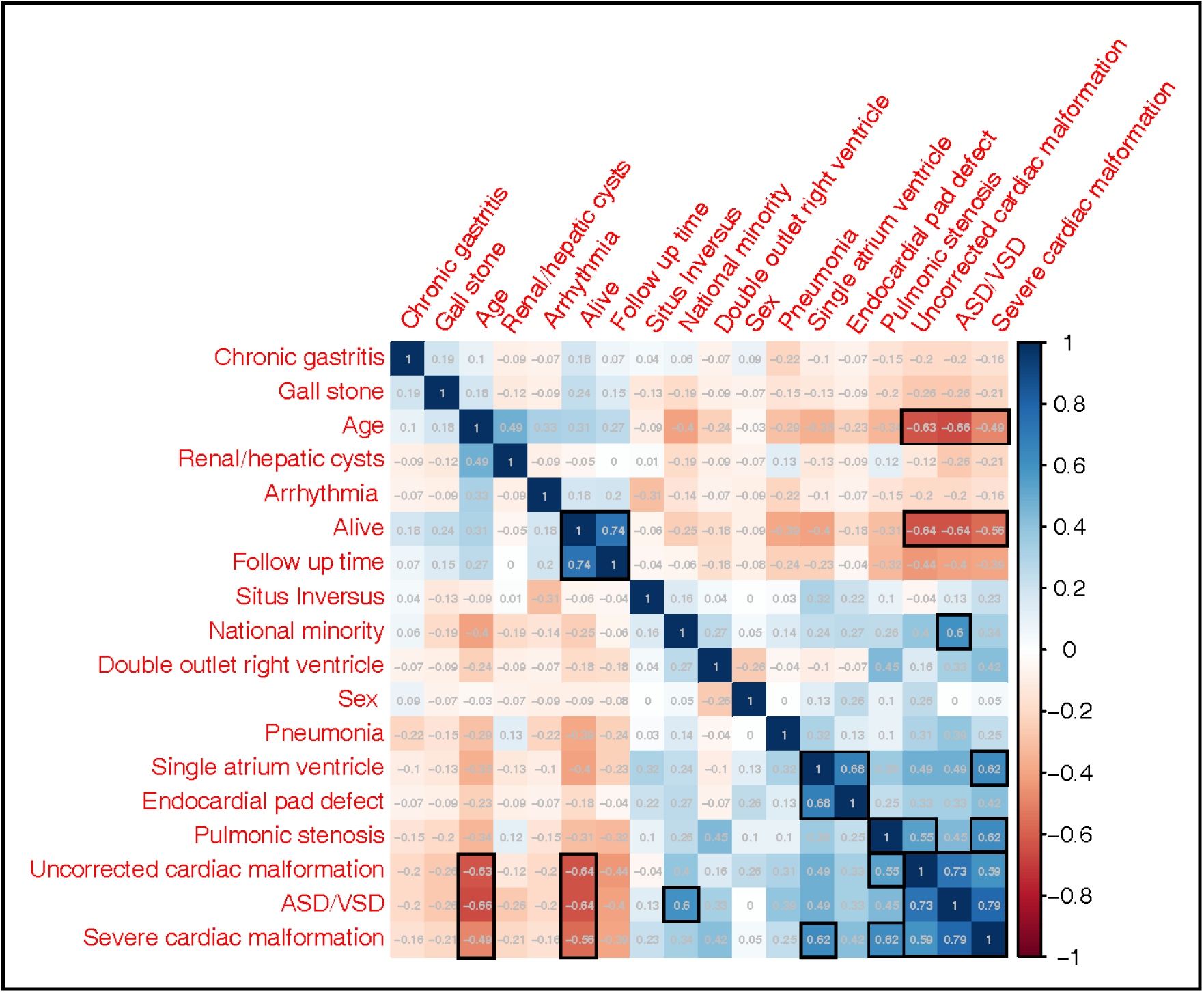
Each cell represents pairwise Pearson correlation for magnitude of correlation through color and r correlation coefficient. Absolute value of correlation coefficient more than 0.5 is represented with black outlined squares. For two-category data, one means yes/male.

#### 3.3.3 Logistic regression analysis

Single-variate analysis suggested that onset age, follow-up time, the incidence rate of cardiac malformation and pneumonia were significantly different in dead group and alive group. According to p-value of variates, clinical importance, scientific knowledge, the incidence rate of uncorrected cardiac malformation, situs inversus and pneumonia were included in the multivariate analysis of survival rate, and forest plot of risk factors for survival was shown in Figure 3. The independent risk factor of survival rate in dextrocardia patients was uncorrected cardiac malformation (OR 22.2391, 95% CI 4.6221∼153.8734, P<0.001).

**Figure 3.**
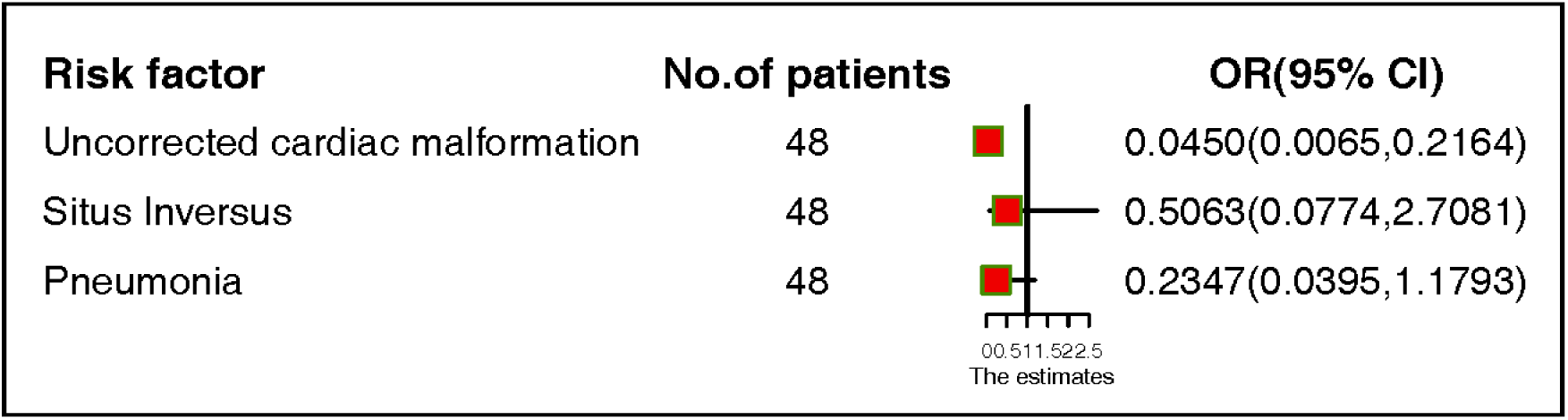
Forest plot of the risk factors for survival rate in dextrocardia patients.

### 3.4 Survival Analysis

The mean follow-up time is 35.46 months. None were lost to follow-up at the end of the study period (May 2020). Among all dextrocardia patients, 18 patients with uncorrected cardiac malformation and 30 patients without cardiac malformation. Kaplan–Meier plot (Figure 4) was shown all death incidents of dextrocardia patients occurred within one year after discharge, and dextrocardia patients’ survival rate was stabilization from one to seven years after discharge. Compared with dextrocardia patients with uncorrected cardiac malformation, those without cardiac malformation (including three patients with corrected cardiac surgery) had higher survival rate (P<0.001). There was no significant difference between healthy cohort and dextrocardia patients without cardiac malformation (P=0.25). 16 death cases of dextrocardia patients included 13 patients (81.25%) with uncorrected cardiac malformation. As for the healthy cohort, only one occurred incident of death. From results above all, we could conclude that uncorrected cardiac malformation is the main cause for death in the dextrocardia patients, and pure dextrocardia with or without situs inversus has no effect on survival rate.

**Figure 4.**
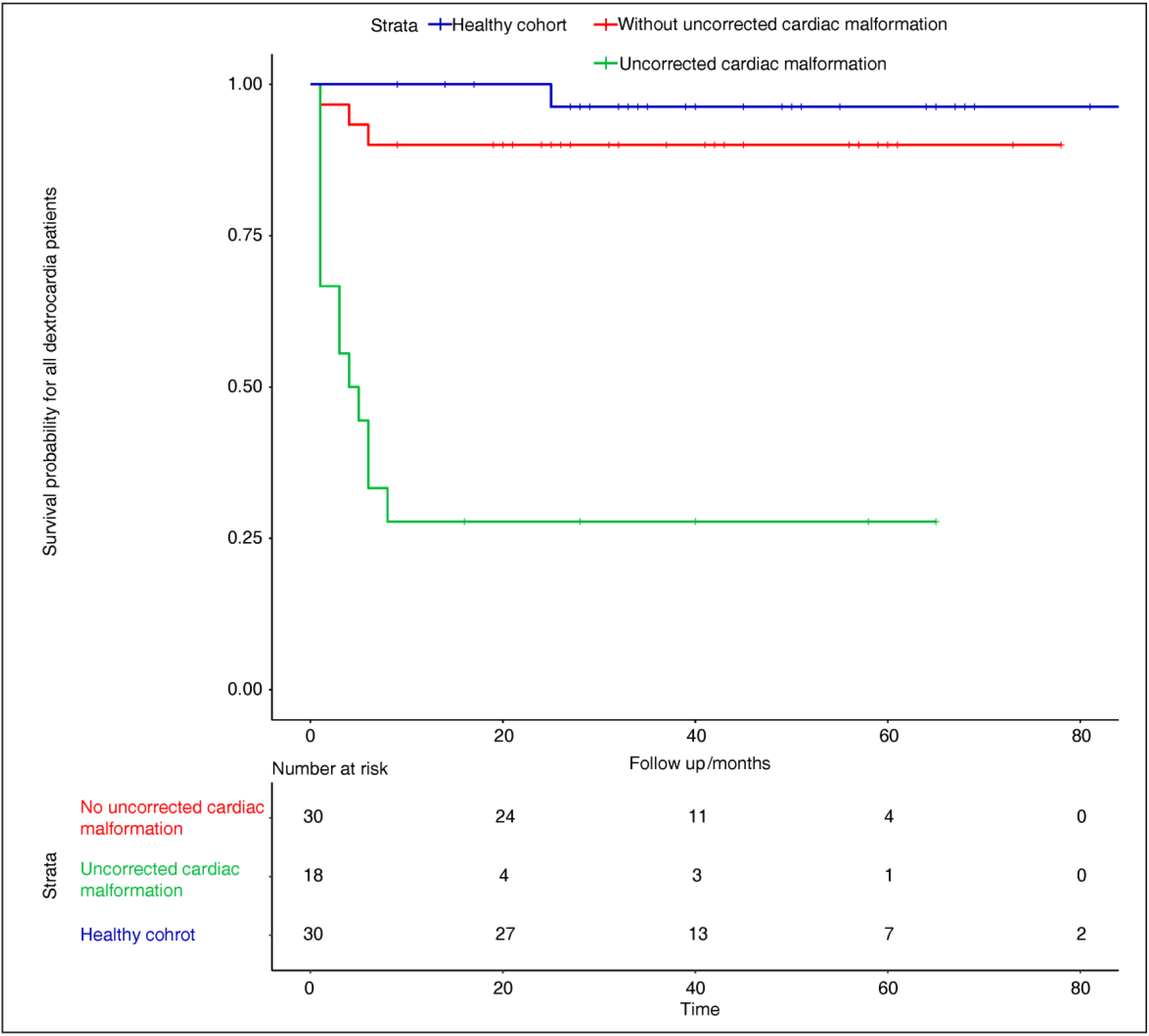
Kaplan–Meier plot showing survival probability over a 7-year time frame.

## 4 Discussion

In our study, the onset age of patients with dextrocardia was bimodal distribution. Less than one year old or more than 50 years old accounted for 62.50% (30/48). The main cause for treatment was pneumonia (18/30). There was no cardiac malformation in the patients older than 50 years old. In contrast, 88.24% patients less than one year old had cardiac malformation (0 vs 88.24%, P<0.001).

Dextrocardia, as a rare congenital anomaly, always has several concomitant malformations, including cardiac malformation, situs inversus, Kartagener syndrome[6], wandering spleen[7], intracranial aneurysm[8], microtia[9]. We also first found dextrocardia patients with medullary sponge kidney and congenital discoid meniscus. Above all concomitant malformations, cardiac malformation and situs inversus are the most common. Northwest China as an undeveloped area, socioeconomic level contributed to the low rate of corrected surgery for dextrocardia patients (6.25%, 3/48). Our study first reported uncorrected cardiac malformation was the major risk factor for dextrocardia patients’ survival time through retrospective analysis.

From the result of correlation analysis, uncorrected cardiac malformation had positive correlation with young onset age, mortality and the rate of minority nationality (r>0.5, P<0.05). Therefore, pneumonia, situs inversus and uncorrected cardiac malformation, as potential risk factors, were incorporated into multivariate logistic regression analysis. The result showed that uncorrected cardiac abnormality is independent risk factor for survival rate of dextrocardia patients.

Kaplan–Meier plot indicated that all death incidents of dextrocardia patients occurred during one year after discharge, and more than three quarters of deaths had uncorrected cardiac malformation. In the 48 dextrocardia patients, only three cases who underwent corrective operation of cardiac deformities, had no major adverse cardiovascular events during follow-up time. Compared to healthy cohort, dextrocardia patients without cardiac malformation had no significant difference of survival rate. Hence, we hold the view that situs inversus didn’t affect dextrocardia patients’ prognosis but cardiac malformation needed for surgical correction as soon as possible.

## 5 Conclusions

Our study confirms the rarity of dextrocardia, which is associated with various malformations besides cardiac malformation. Uncorrected cardiac malformation is the major death factor of dextrocardia patients, and dextrocardia combines only situs inversus has no effect on patients’ survival.

## Data Availability

The data used to support the findings of this study are available from the first author upon request.

